# Age-based risk estimates for *C9orf72*^RE^-related diseases: Theoretical developments and added value for genetic counseling

**DOI:** 10.1101/2025.08.08.25333050

**Authors:** Dominique de Vienne, Damien M. de Vienne

## Abstract

The *C9orf72* hexanucleotide repeat expansion is the most common genetic cause of amyotrophic lateral sclerosis (ALS) or frontotemporal dementia (FTD). In genetic counseling, children of mutation carriers are often told that they have a 50% risk of carrying the mutation, but this figure does not take into account the fact that penetrance is age-related, with a unimodal distribution of disease onset around 58 years of age. Using a Bayesian approach, we developed a theory to calculate the probability of carrying the mutation for asymptomatic relatives (children/siblings and grandchildren/niblings) as well as the probability of developing ALS/FDT within a given time frame, based on their age. Using published data on age-related penetrance, we then calculated these probabilities and developed an online simulator that makes it easy to calculate them on a case-by-case basis. The conditional probabilities obtained can be very different from Mendelian values. For example, a 70-year-old asymptomatic child born to a carrier has approximately a 6% risk of being a carrier, which is far from 50%. For grandchildren, taking into account both their age and that of their parents also leads to figures that are much lower than those obtained if only their age were considered. For consultands, the decision to undergo testing is based in part on risk estimates. In this regard, the refined estimates and simulator we propose may prove to be valuable tools for genetic counseling for families affected by ALS/FTD linked to the *C9orf72*^RE^ mutation. In addition, the formulas used in this study could also be used to calculate risk estimates for other diseases caused by autosomal dominant mutations with age-dependent penetrance.

**Author Summary:** Amyotrophic lateral sclerosis and frontotemporal dementia are often linked to a mutation called *C9orf72* repeat expansion. Relatives of mutation carriers are usually informed of a 50% (children) or 25% (grandchildren) inheritance risk. These values, however, ignore the age of the relatives, although the risk is strongly age-dependent for these diseases. We developed the necessary mathematical framework to compute these risks, and we applied these developments using a dataset relating to the age of onset of the disease. Our results show that actual risks are often far lower than the standard probabilities given to consultands. For instance, a 70-year-old child of a carrier has an estimated 6% probability of carrying the mutation, rather than 50%. To facilitate clinical use, we offer an online tool that provides instant age-adjusted risk estimates. These findings enable more accurate genetic counseling and promote informed decision-making regarding genetic testing.

## Introduction

Amyotrophic lateral sclerosis (ALS) and frontotemporal dementia (FTD) are two neurodegenerative diseases primarily characterized by motor and behavioral dysfunction, respectively [22, 11, 16]. Both conditions have been genetically linked to the mutation *C9orf72*^RE^, a hexanucleotide repeat expansion in the *C9orf72* gene, which is the most common genetic cause of ALS (34.2%) and FTD (25.2% of the cases [7, 23, 1, 3]). This mutation being autosomal and dominant, the children and siblings of an affected individual have, *a priori*, 50% risk of being carrier of *C9orf72*^RE^, whereas the grandchildren and niblings (nieces and nephews) have a 25% risk. Molecular genetic testing can confirm the genetic origin of ALS/FTD in affected individuals, and can also be used to identify carriers of *C9orf72*^RE^ among relatives. However, the choice for relatives to be tested or not is far from trivial, and should be based not only on a thorough clinical assessment [13], but also on a precise estimate of the risks.

The standard probability of 0.5 and 0.25 assigned respectively to children/siblings and grandchildren/niblings eligible for testing is a marginal probability that does not take into account the age of the consultands. However, the penetrance is clearly age-dependent, with a distribution of the age of onset of symptoms centred around 58 years [13, 18], so that the probability of unaffected individuals to be carrier of *C9orf72*^RE^ decreases with age, and the probability of developing the disease in the future drops rapidly once the median age of symptom onset has been exceeded.

In the case of cancers, well-established theoretical frameworks [21, 6, 14] along with online platforms (CanRisk [5], ASK2ME [4]) allow estimation of age-dependent carrier probabilities and future disease risks. These approaches rely on a general Bayesian framework that is, in principle, applicable beyond oncology. These approaches rely on a general Bayesian framework that is, in principle, applicable beyond oncology. However, to our knowledge, no applications of these tools have been proposed for neurodegenerative diseases such as ALS/FTD, despite their significant implications for human health. Therefore, there is a need for dedicated theoretical developments for *C9orf72*^RE^ and other autosomal dominant mutations, together with a handy online tool for age-dependent risk estimation, to provide ALS/FTD genetic counseling with a level of quantitative information comparable to that available in oncology.

In this article, we use a Bayesian approach to estimate conditional, age-specific probabilities of carrying a mutation and developing the associated disease later in life for the relatives of a carrier of an autosomal dominant mutation. We derived these probabilities in the case of both complete penetrance of *C9orf72*^RE^ with age and incomplete penetrance, *i*.*e*., when a fraction of carriers is assumed to never develop the disease.

We applied these developments to the data on age-related penetrance of *C9orf72*^RE^ from the large cohort of Murphy et al. [18], and provided numerical estimates of these probabilities. Our results show that the risks of being a carrier of the variant and of developing the disease over time can deviate in a large extent from *a priori* Mendelian marginal probabilities usually given to the consultands. For example, in the case of complete penetrance with age, a child presenting no symptoms at the age of 70 would have a risk of about 6% of being a carrier of *C9orf72*^RE^ (instead of 50%) and a 5.5% risk of developing the disease in the following 10 years. If 20% of carriers are non-penetrant, the risk of being carrier rises to about 20%, still well below 50%, but the risk of developing the disease in the next 10 years falls to about 4%.

Similarly to established platforms such as CanRisk and ASK2ME, which are dedicated to cancer risk estimation, we have integrated our results into a practical online simulator designed for genetic counselors, providing reliable age-related risk estimates for ALS/FTD. This can help them meet the expectations of consultands who need as much information as possible to decide whether they wish to undergo testing for *C9orf72*^RE^. The methodology presented could also be applied to any disease caused by autosomal dominant mutations, after implementation of available age-related penetrance data.

## Description of the method

We addressed two linked questions regarding the unaffected relatives of carriers of autosomal dominant mutations that have age-dependent penetrance: (i) what is the probability of being carrier of the mutation? (ii) what is the probability of developing the disease over the next *n* years? We considered either that penetrance tends towards 1 with age, or that there is a fraction *x* of carriers who never develop symptoms, *i*.*e*., who are “non-penetrant” carriers, a supposedly non-heritable characteristic. (see Discussion). The equations we obtained using a Bayesian framework are summarized in Table 1, which offers a comprehensive overview of the probabilities useful in genetic counseling for the children and grandchildren of a mutation carrier (see detailed mathematical developments in S1 Appendix). For simplicity, we only mention below children and grandchildren, but the results apply as such to siblings and niblings, respectively, because they have the same Mendelian probabilities of inheriting the mutation. Of the eight equations of Table 1, only equations 2 and 8 had previously been derived explicitly [27, 19, 20], although they all fall within the same general Bayesian framework. The probabilities are quite simple functions of the age-related penetrance *π*_*t*_ (or 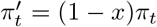), with a similar structure of the equations in columns ‘Child’ and ‘Grandchild’: there is just one additional term 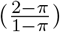 in the equations relating to grandchildren, which is the inverse of the probability of being a carrier for the child 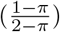.

**Table 1:**
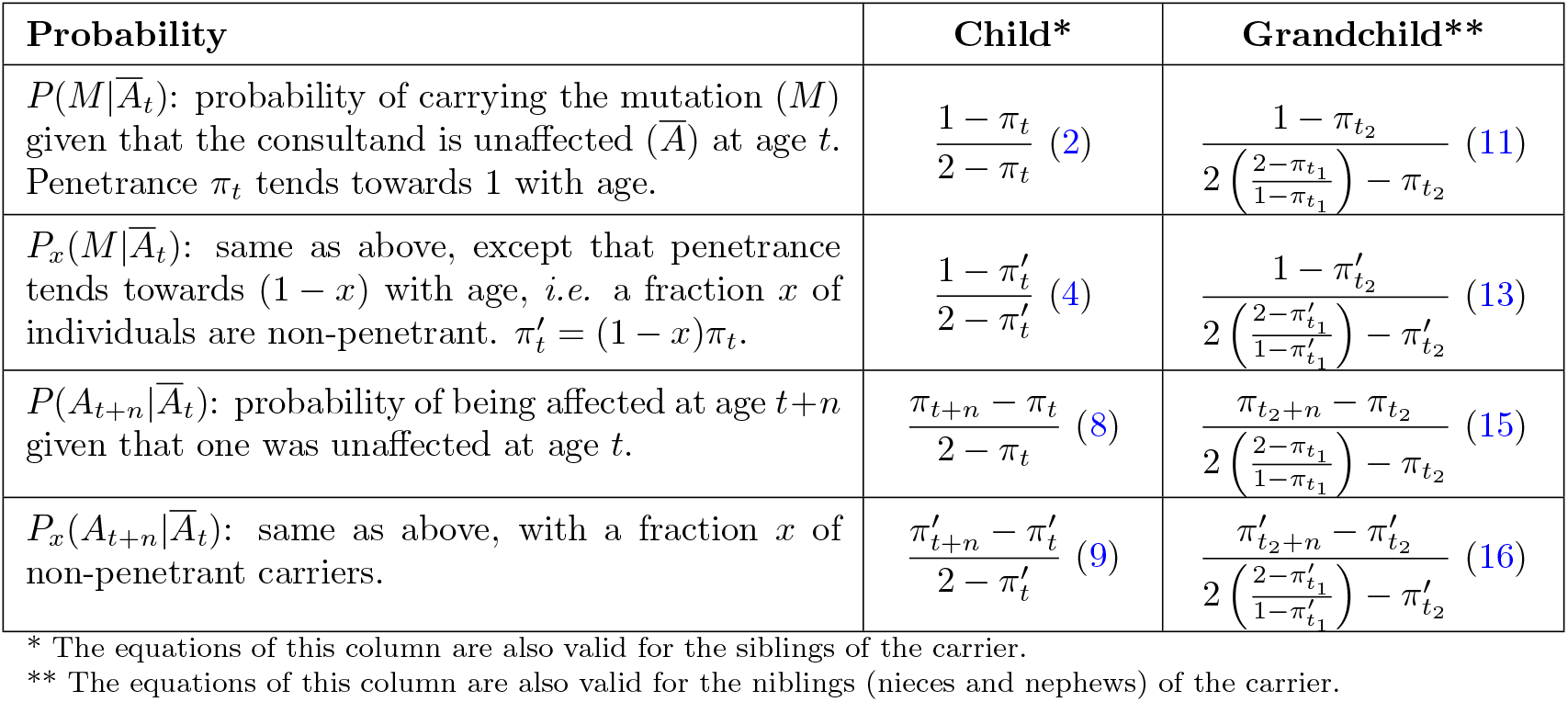
Formulae to be used for computing the risk probabilities for non-affected relatives of a carrier of an autosomal dominant mutation. The penetrance at age *t* is denoted *π*_*t*_. 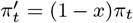 is the penetrance at age *t* when a fraction *x* of the carriers never get affected. *t*_1_ and *t*_2_ are the ages of the child and the grandchild, respectively. The numbers in parentheses correspond to the equation numbers in S1 Appendix.

## Applications

### Probability estimates from empirical data on *C9orf72*^RE^ penetrance

#### Dataset description

Using the equations presented in Table 1, we computed the age-dependent probabilities of (i) carrying the variant *C9orf72*^RE^ and (ii) developing ALS/FTD symptoms, taking advantage of the age-dependent penetrance data from [18] that are based on a cohort of 1169 individuals carrying *C9orf72*^RE^. Among these individuals, 1146 had FTD and/or ALS symptoms, with age at symptom onset varying from 25 to 83 (**Figure 1A**), and 23 were asymptomatic. We discarded these asymptomatic cases as they do not inform us on the actual fraction of non-penetrant carriers in the population, for two reasons: first, a certain proportion of them, if not all, may end up developing the disease; second, if they are real non-penetrant carriers, their number might be underestimated as healthy individuals are less likely to consult and undergo genetic testing. Using the 1146 symptomatic individuals we got an estimate of the age-related penetrance *π*_*t*_ by dividing the cumulative count at each age by the total number of individuals (**Figure 1B**).

**Figure 1:**
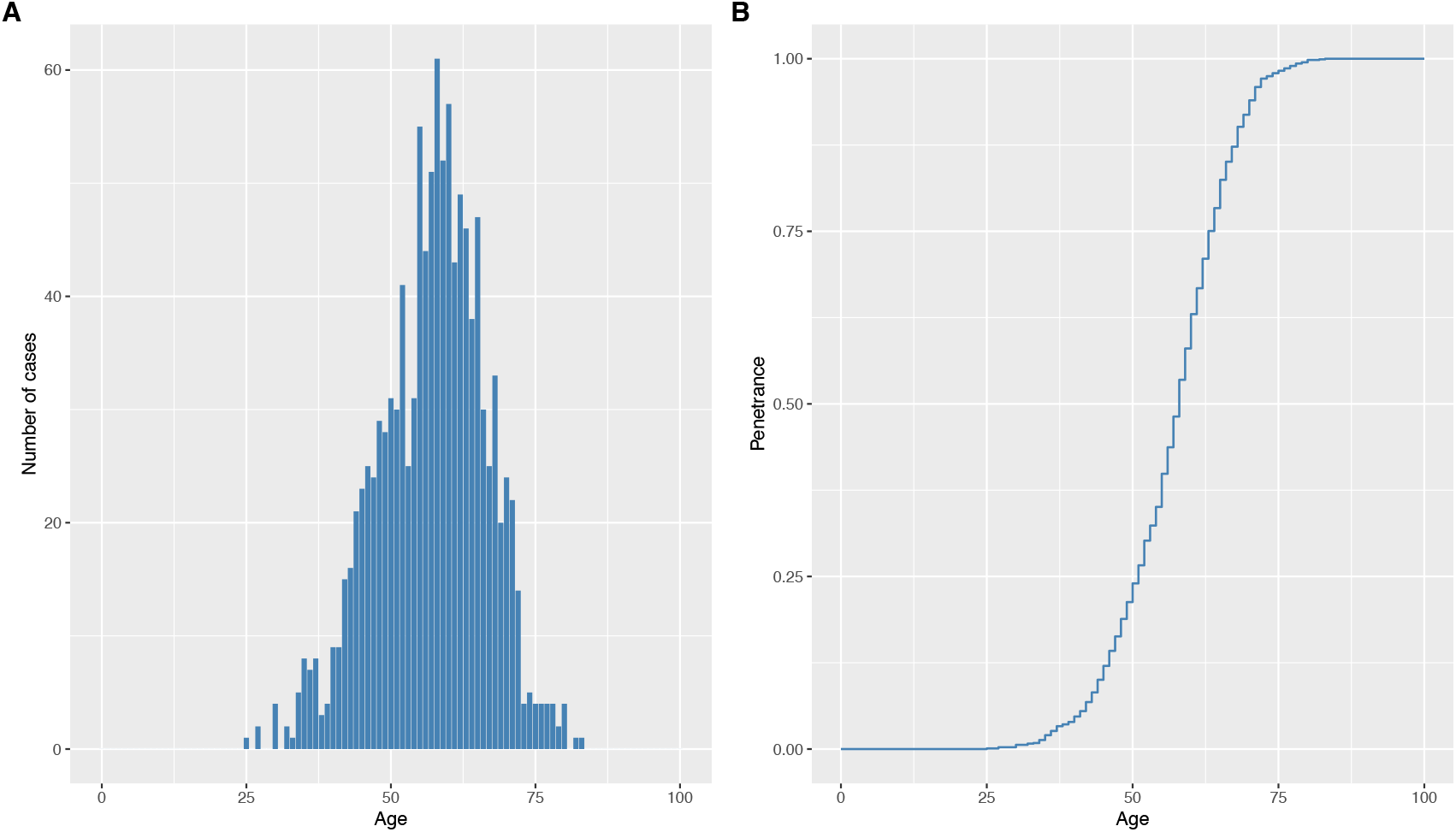
Distribution of age at onset of symptoms in 1146 individuals carrying the *C9orf72*^RE^ mutation (**A**) and corresponding age-dependent penetrance of the mutation (**B**). Data from [18].

#### Age-dependent probability of carrying *C9orf72* ^**RE**^

We used equation 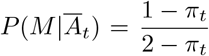 (equation 2 in Table 1) to estimate the probability of asymptomatic children to carry the mutation, given their age (**Figure 2A**, dark blue curve). This estimate is very close to 0.5 at age 25-30, then decreases to 0.27 at age 60 and 0.06 at age 70. So the risk of carrying the variant for healthy individuals is very dependent on their age and can be far from the 50% risk commonly communicated to the consultands for this disease (**Figure 2A**, dashed grey line).

**Figure 2:**
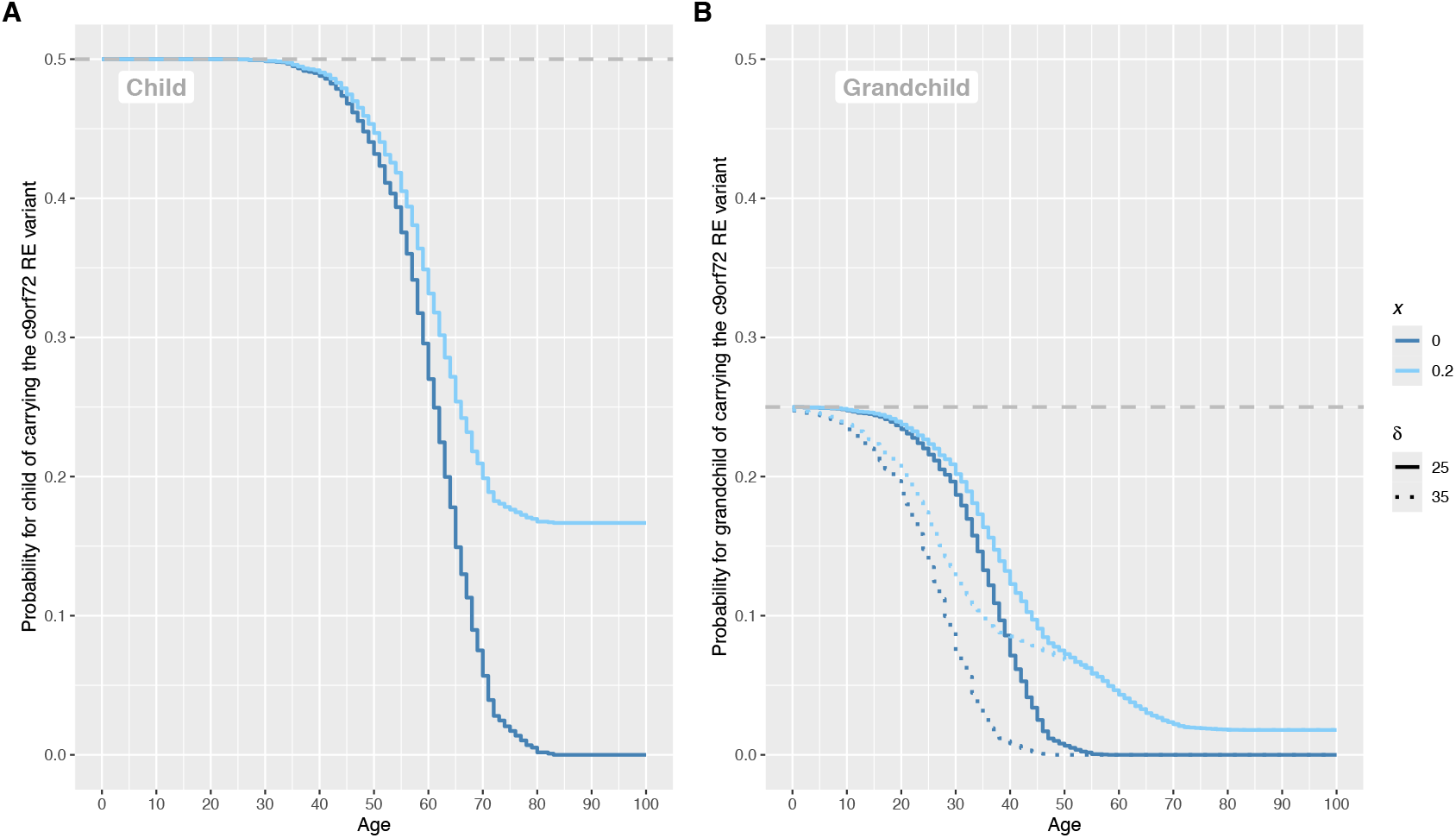
Age-related risk estimates for asymptomatic child and grandchild of a carrier of the *C9orf72*^RE^ mutation. **A.** Relationship between age and the probability of being carrier for an asymptomatic child, assuming that the fraction of non-penetrant carriers is either *x* = 0 (dark blue curve) or *x* = 0.2 (light blue curve). **B**. Same relationships for a grandchild of the carrier, in the case where the age difference between the child and the grandchild is 25 years (solid lines) or 35 years (dotted curves). Color codes: same as in **A**.

If a fraction *x* of the population is non affected by *C9orf72*^RE^ irrespective of age, the probability of carrying *C9orf72*^RE^ is given by equation 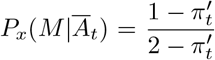 (equation 4) (**Figure 2A**, light blue curve). This probability increases with the fraction of non-penetrant carriers, reaching the maximum probability of 0.5, regardless of age, in the unrealistic assumption where 100% of carriers were non-penetrant (*x* = 1). This reflects the fact that being unaffected at a given age is less and less informative as *x* increases.

These probabilities can also be calculated for the carrier’s grandchildren, when their parent (the carrier’s child) is not affected and their carrier status is unknown. If *x* = 0, the probability of being carrier is 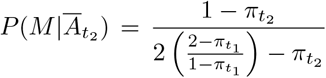 (equation 11), which depends on both the age of the grandchildren and the age of their parent (**Figure 2B**, dark blue curve). For instance, if a grandchild is 45 and its parent (child of the carrier) is 70 (*i*.*e. δ*, the age difference between both, is 25 years), the probability is 0.025, a value ten times lower than the prior probability of 0.25 for grandchildren to carry *C9orf72*^RE^ in the absence of information regarding the age-dependent penetrance.

If *x* > 0, this probability is 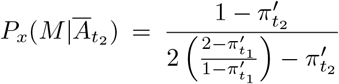 (equation 13). With *x* = 0.2, the probability for the same 45-year old grandchild rises to 0.09 (**Figure 2B**, light blue curve). Indeed, for the reason mentioned above, the larger the *x*, the higher the probability of being a carrier. Consistently, in the theoretical case where *x* would be equal to 1 (*i*.*e*. the penetrance is null), the limit would be 0.25, which is the prior Mendelian probability of being carrier for a grandchild (**Figure 2B**, dashed grey line).

If the age difference between the child and the grandchild is higher, the probability curves are shifted to the left. The probability of carrying the variant is lower since the unaffected child is older and has a lower probability of being a carrier. For example, with *δ* = 35 years, the 45-year old grandchild has a probability of being carrier of 0.07% if *x* = 0 and 7.6% if *x* = 0.2 (**Figure 2B**, dark and light blue dotted curves), to be compared to 2.5% and 9.1%, respectively, when *δ* = 25 years (**Figure 2B**, dark and light blue solid curves).

#### Age-dependent probability of developing the disease at a later age

The probability that a child aged *t* years will develop the disease over the next *n* years is 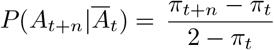 (equation 8). By setting *n* = 10 years, we observe that the probability is null up to age 20, then increases to reach a maximum of 0.26 at age 55, and then drops to 0 at age 80 (**Figure 3A**, red curve). If we consider a longer-term risk, for example for *n* = 20 years (**Figure 3C**, red curve), the maximum probability of declaring the disease is higher (0.4) and occurs for a younger age (51 years). Note that as *n* increases, the probability tends towards 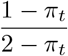, the probability of carrying the variant, which is consistent.

**Figure 3:**
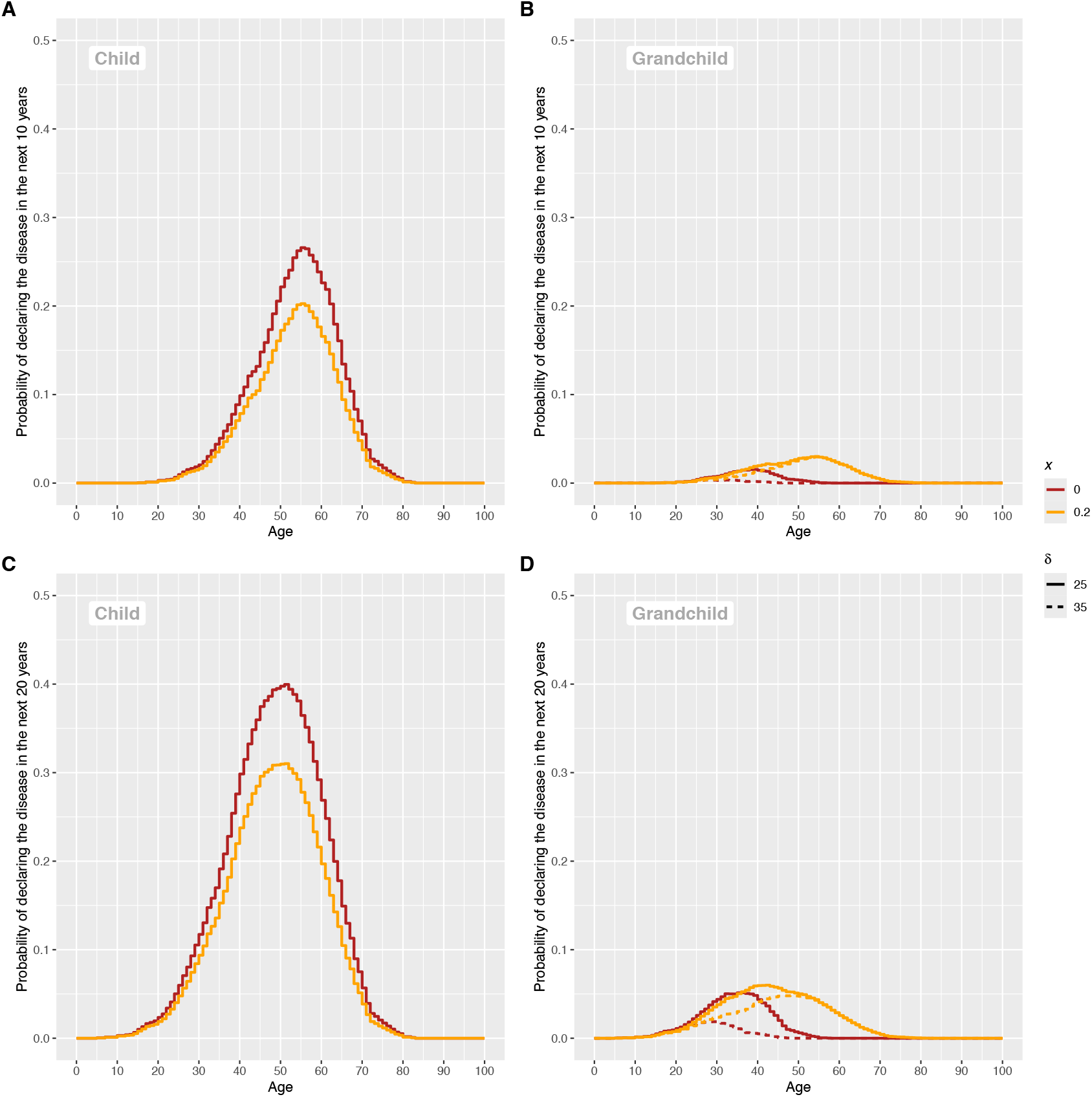
Age-related estimates for asymptomatic children (**A** and **C**) and grandchildren (**B** and **D**) of the risk of developing the disease in the next *n* = 10 years (**A** and **B**) or *n* = 20 years (**C** and **D**). In each plot, the relationship between the age and the probability to develop the disease is displayed for a fraction of non-penetrant carriers equal to *x* = 0 (red curve) or *x* = 0.2 (orange curve). For the grandchild risk estimates, the relationship is given for two different age differences to their relative (child of the carrier) : *δ* = 25 years (solid lines) or *δ* = 35 years (dotted lines).

If the penetrance is incomplete, the probability of declaring the disease is 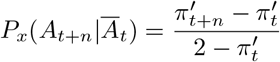 (equation 9). The derivative of this function with respect to *x* is negative. So, whatever *π*_*t*+*n*_ and *π*_*t*_, the probability of declaring the disease decreases as *x* increases (**Figure 3A** and 3C, orange curves), even though the probability of carrying *C9orf72*^RE^ increases.

For the grandchildren, we have 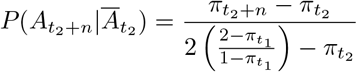 (equation 15), a probability much lower than that calculated for children at *n* = 10 years (**Figure 3B**, red curve). When *n* increases, this probability tends towards 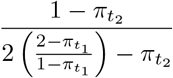, the probability of carrying *C9orf72*^RE^. If *x* > 0, we have 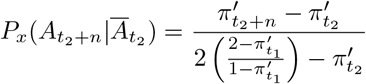 (equation 16), which tends towards 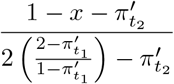 as *n* increases.

Equation 16 has a negative second derivative with respect to *x*, meaning that there is a value of *x* that maximizes the probability of developing the disease. We show in S1 Appendix that this value, noted *x*_*m*_, only depends on 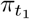, the age-dependent penetrance of the child. When 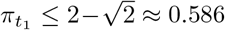, corresponding to 59 years of age or younger, *x*_*m*_ = 0. The fact that the probability of declaring the disease for a grandchild is the highest when the penetrance is complete was expected. However, when 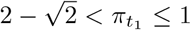, we show that *x*_*m*_ > 0, with the upper limit 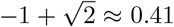. This means that the probability of declaring the disease is higher when there are non-penetrant carriers. This seemingly paradoxical result can be understood in the following way. Given that the children concerned have adult descendants, they are necessarily relatively elderly individuals. When *x* = 0, such unaffected child are very likely not carrier of *C9orf72*^RE^, so the probability that their descendants (grandchildren) will develop the disease is negligible. When *x* > 0, an elderly person may not be affected either because they are not a carrier or because they are a non-penetrant carrier. In the latter case, grandchildren may inherit *C9orf72*^RE^ and develop the disease if they belong to the (1 − *x*) fraction of carriers. In any case, we see that the probability of developing the disease remains quite low for the grandchildren whose parent is unaffected (**Figure 3B** and D, orange curves).

### An online simulator for genetic counseling

Based on the equations presented in Table 1 and Murphy et al.’s data [18], we developed an online simulator to easily estimate the risks of (i) carrying the *C9orf72*^RE^ variant and (ii) developing the disease (ALS or DFT) in the next *n* years. This tool, available free of charge online at https://lbbe-shiny.univ-lyon1.fr/ftd-als/, is designed to facilitate genetic counseling provided by professionals to consultands, with an interactive and intuitive way to compute the risk estimates. The tool is organised in three panels (**Figure 4**). In the left-hand panel, the input parameters can be set: age of the consultand, relationship to the affected individual (child/sibling or grandchild/nibling), age, age of its parent (when the consultand is a grandchild or a nibling), assumed disease penetrance, and time frame to compute the risk of declaring the disease. In the middle panel, the distributions of the probability to carry *C9orf72*^RE^ and the probability to declare the disease are plotted as a function of age, and the age of the consultand is highlighted by a dotted vertical line. In the right-hand panel, a summary of all the information is provided, along with the option to download a one-page pdf file.

**Figure 4:**
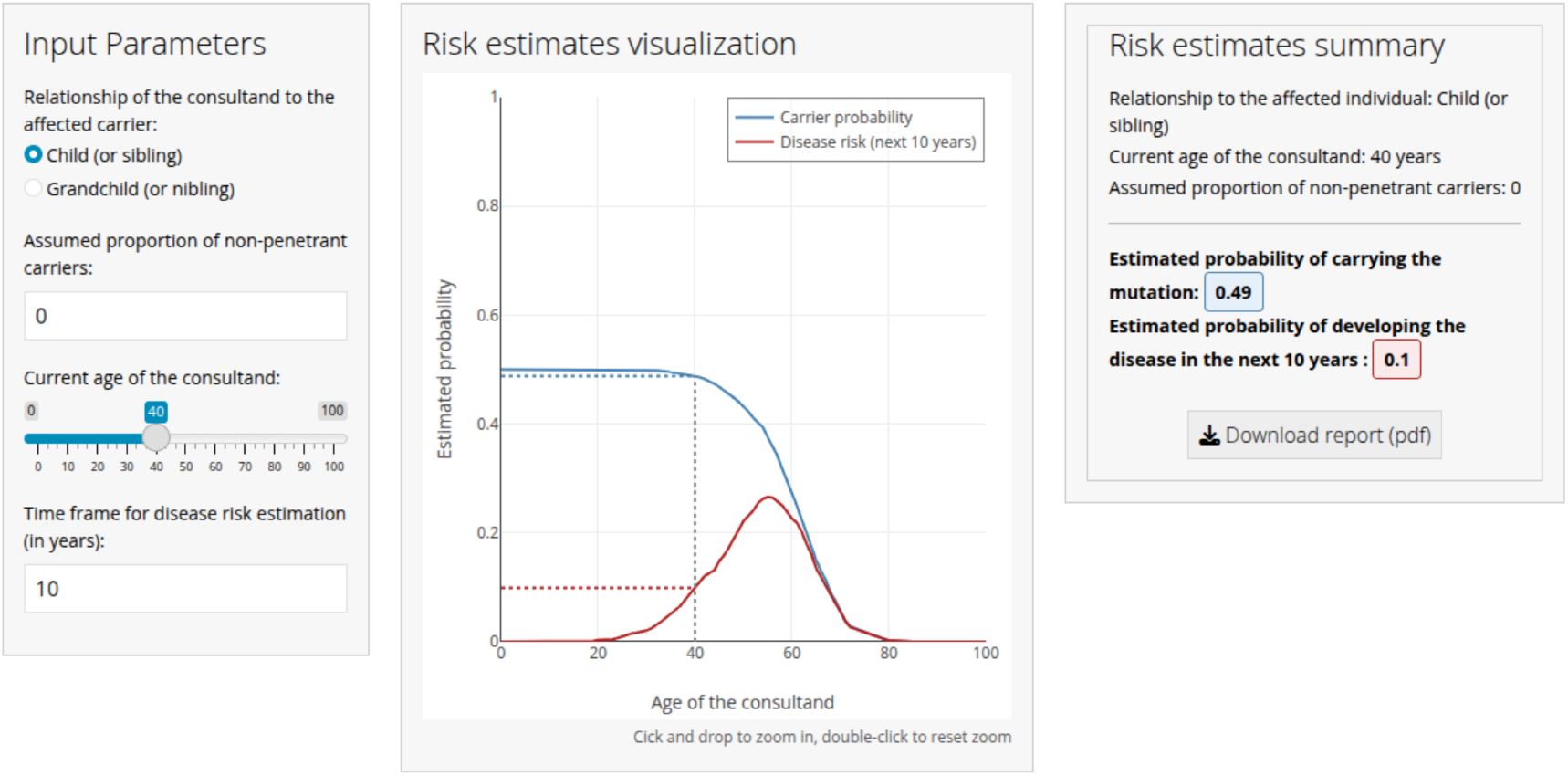
Interface of the web-based tool for genetic counseling for the *C9orf72*^RE^-related ALS/FTD diseases, available at https://lbbe-shiny.univ-lyon1.fr/ftd-als/. The application includes three panels. Left: input of consultand’s information; Center: graphical visualization of the age-dependent probability of being carrier and of the disease risk; Right: summary of results with the option to download a report in PDF format.

## Discussion

Risk assessment for diseases such as ALS/FTD faces two levels of uncertainty: genetic randomness and extreme variability in the age of onset of symptoms. For this reason, it is mainly the clinical and psychological aspects that are emphasized in genetic counseling [2, 24]. Most often, the only figures communicated to unaffected relatives of mutation carriers are the Mendelian-based risks of being carrier, *i*.*e*., 50% for children and siblings, and 25% for grandchildren and niblings. However, these figures may differ significantly from the risk estimated when taking into account the age of the consultands. Since risk value is one of the key factors in the decision to get tested, it is important to provide the consultands with values that are as reliable as possible.

In this study, we addressed two questions psychologically crucial for unaffected relatives of a carrier of a deleterious mutation: given their age, what is their probability of being a carrier of *C9orf72*^RE^, and what is their probability of developing the disease in the future? While there are quite a few articles on penetrance estimation from family history of diseases or from cohort analyses [27, 20, 25, 9, 26], few studies formally address these issues [27, 20, 19]. In addition, these formal studies, which have remained quite confidential, provide only partial and disparate answers and do not result in handy guides for genetic counseling. Our work could be viewed as a special case of the general Bayesian framework already used in medical genetics, particularly for cancer risk assessment. However, its value lies in the fact that the risk estimates are easily accessible via a simulator and directly applicable to genetic counseling regarding ALS/FTD associated with the *C9orf72*^RE^ variant.

We have formalized how to use the distribution of age at symptom onset of ALS/FTD caused by *C9orf72*^RE^ to estimate the risk of being a carrier and developing the disease for the children/siblings and grandchildren/niblings of a carrier, considering that penetrance tends toward either 1 or a limit. The eight equations we have established (two of which were proposed independently [19, 27]) have been implemented in an online simulation tool that allows the desired probabilities to be easily estimated.

The age-dependent penetrance values we used come from the large Murphy et al’s cohort [18]. We are aware that such empirical data suffers from various biases, such as the under-representation of aged people and the fact that the vast majority of patients are European, North American or Australian. However, moderate population-specific founder effects appear to exist for this mutation [12], and the penetrance estimates are roughly similar in other cohorts [13], even in those including a fraction of patients from non-European origin [15, 17]. Various refinements for estimating the penetrance could be considered, such as taking into account the family history of the disease [27, 25, 9] or the intra-family correlation for the onset of the disease [10], but we do not know whether this is worthwhile, given the uncertainty associated with the inevitable limited size of the families. In any case, if new approaches for penetrance estimate lead to more reliable values [28], the simulator can and will be updated.

The risk estimates we found can be much lower than the Mendelian values classically given to the consultands. For example, even for children whose median age of symptom onset is 58 years, the probability of carrying the variant is 0.32 and not 0.5, and drops at 0.1 at 67-68 years. For grandchildren, taking parental status into account can make a big difference. For example, if the child is 70 and the grandchild 45, the probability of the grandchild carrying *C9orf72*^RE^ is only 0.025 (2.5% risk), whereas it is almost ten times higher (0.23) if parental status is ignored.

The existence of a fraction *x* of non-penetrant carriers is assumed [18, 8], but its value is not known. Environmental, genetic (epistasis) and demographic (age at death) factors combine to make its estimate methodologically out of reach, or at best extremely imprecise, and non-penetrant carriers can have affected descendants. However, introducing *x* into the calculations makes it possible to analyze the impact of the presence of non-penetrant carriers on the calculated risks. For illustrative purpose, we set *x* at 0.2, the upper limit proposed by Murphy *et al*. [18], but the whole range of risks can be explored by varying *x*. We believe that this may be of interest to counselors and consultands.

Consultands who wish to undergo presymptomatic testing must be fully informed, both on genetic and psychological issues, and be given sufficient time to determine whether they are ready to receive the results of a genetic test [24]. In this context, we show that consultands whose age is beyond the median age of onset of symptoms have a risk of being carriers, and hence of being affected, markedly below the Mendelian proportions. If the consultand is aged 40 or 50, the difference between the marginal probability and the conditional probability is moderate. However, since the *C9orf72*^RE^ variant is generally detected in symptomatic patients of advanced age, their relatives of the same generation (such as the siblings and first cousins) may wish to seek genetic counseling. For these individuals, the use of conditional probabilities has a significant effect on risk estimates.

We think that our results can represent a useful contribution to genetic counseling. Firstly, because many individuals express the sentiment that the anxiety of living with the unknown is worse than knowing whether or not they are at genetic risk [2]. Secondly, because this is important information in cases of disagreement between generations, particularly when the desire to have children prompts the consultands to ask their parent to be tested.

Finally, we emphasize that our approach is valid for any other disease caused by an autosomal dominant mutation, provided empirical distributions of the age of onset of symptoms is available. This broad applicability makes our framework a generalizable tool for genetic counseling and risk assessment.

## Standard Protocol Approvals, Registrations, and Patient Consents

This study uses public data from an anonymous patient cohort [18]. It does not involve any experimental work or clinical trials, but only probabilistic developments and their application to a widely used dataset. Therefore, the issue of patient consents or ethics committee approval does not arise.

## Data availability

All the code used for data analysis, for figure preparation, and the source code of the online “shiny” application is available at https://github.com/damiendevienne/ftd-als/

## Acknowledgments

We are very grateful to Dr. Mathieu Fauvernier for mathematical advice on a preliminary version of the manuscript, and to three reviewers for their helpful suggestions.

