## Supplementary information for "Age-based risk estimates for *C9orf72*^RE^-related diseases: Theoretical developments and added value for genetic counseling"

### S1 Appendix

#### Theoretical developments underlying age-based risk estimates for relatives of individuals carrying *C9orf72*$^{\mathrm{RE}}$

***Note**: The following developments consider the case of unaffected children and grandchildren of a mutation carrier. They are equally valid in the case of siblings and niblings, respectively, because the probabilities of having inherited the mutation from the carrier are the same, i.e.* 0.5 *and* 0.25.

### 1 Probability of carrying the mutation for a child of the carrier

#### 1.1 Case 1: The penetrance tends towards 1 with age

The probability that an individual with one parent carrying the *C9orf72*$^{\mathrm{RE}}$ mutation is also a carrier is

$$P(M) = \frac{1}{2}$$

where $M$ stands for 'mutation'. If we know that this individual has no symptoms of the disease at age $t$, this probability writes:

$$P(M|\overline{A}_t),$$

where $\overline{A}_t$ stands for 'unaffected at age $t$'. This probability can be estimated by applying the Bayes' theorem:

$$P(M|\overline{A}_t) = \frac{P(\overline{A}_t|M)P(M)}{P(\overline{A}_t)} \tag{1}$$

where

- $P(\overline{A}_t|M)$ is the probability of not being affected at age $t$ despite being a carrier of the mutation. Note that $1 - P(\overline{A}_t|M) = P(A_t|M)$ is the age-related penetrance, which we will refer to as $\pi_t$ hereafter. We assume here that there is no non-penetrant carriers, *i.e.* that penetrance $\pi_t$ tends towards 1 with age.
- $P(\overline{A}_t) = 1 - P(A_t)$ is the probability of not being affected at age $t$. $P(A_t)$ is the product of the probability of carrying the mutation and of the penetrance: $P(A_t) = P(M)\pi_t$.

Therefore equation 1 can be rewritten in terms of age-related penetrance:

$$P(M|\overline{A}_t) = \frac{1 - \pi_t}{2 - \pi_t} \tag{2}$$

This equation expresses the age-related probability for an unaffected child to carry the mutation (see also [8]).

##### 1.2 Case 2: There is a proportion $x$ of non-penetrant carriers

It may happen that individuals carrying the mutation never get affected, regardless their age [3, 4, 5, 7]. They are called non-penetrant carriers. Their presence alters both the probability of carrying the mutation for asymptomatic individuals and the probability of developing the disease.

Let $x$ the proportion of non-penetrant carriers. The probability that an unaffected individual of age $t$ is a carrier of the mutation writes (equation 1)

$$P_x(M|\overline{A}_t) = \frac{P_x(\overline{A}_t|M)P(M)}{P_x(\overline{A}_t)} \tag{3}$$

where

- The index $x$ of $P$ means $x > 0$.
- $P_x(\overline{A}_t|M) = 1 - P_x(A_t|M)$. As only the fraction $1 - x$ of the carriers can be affected, the penetrance $P_x(A_t|M) = \pi'_t$ is equal to $(1 - x)\pi_t$. Because $\pi_t$ tends towards 1 with age, $\pi'_t$ tends towards $1 - x$.
- $P_x(\overline{A}_t) = 1 - P_x(A_t)$. The conditions for being affected are to be a carrier of the mutation and not to be a non-penetrant carrier. So $P_x(A_t) = P(M)(1 - x)\pi_t$.

Introducing these terms in equation 3, we get

$$P_x(M|\overline{A}_t) = \frac{1 - \pi'_t}{2 - \pi'_t} \tag{4}$$

The derivative of $P_x(M|\overline{A}_t)$ with respect to $x$ is positive, meaning that the probability of being carrier of the mutation when there is no symptom at age $t$ increases with $x$. This is due to the fact that a larger number of individuals may be asymptomatic despite being carriers. At the limit, if $x = 1$, $P_x(M|\overline{A}_t) = \frac{1}{2}$.

#### 2 Probability of developing the disease later in life for a child of the carrier

An important issue for individuals at risk is not only the probability of carrying the mutation as seen above, but also the probability of developing the disease later in life, *e.g.* in the next $n$ years. In the following developments, we have neglected the risk of ALS or FTD unrelated to *C9orf72*$^{\mathrm{RE}}$, which is relatively very low: the lifetime risk is only 1/350-400 for ALS [1] and about 1/700 for FTD [2].

#### 2.1 Case 1: The penetrance tends towards $1$ with age

For a nonaffected carrier of the mutation at age $t$, the probability of being affected at age $t+n$ writes

$$P_M(A_{t+n}|\overline{A}_t) = 1 - P_M(\overline{A}_{t+n}|\overline{A}_t)$$

where $P_M$ stands for the probability in the presence of the mutation. From Bayes' theorem, we have

$$P_M(\overline{A}_{t+n}|\overline{A}_t) = \frac{P_M(\overline{A}_t|\overline{A}_{t+n})P_M(\overline{A}_{t+n})}{P_M(\overline{A}_t)}$$

Because $P_M(\overline{A}_t|\overline{A}_{t+n}) = 1$, it comes

$$P_M(A_{t+n}|\overline{A}_t) = 1 - \frac{P_M(\overline{A}_{t+n})}{P_M(\overline{A}_t)} \tag{5}$$

So (see also [6]):

$$\begin{aligned} P_M(A_{t+n}|\overline{A}_t) &= 1 - \frac{1-\pi_{t+n}}{1-\pi_t} \\ &= \frac{\pi_{t+n}-\pi_t}{1-\pi_t} \end{aligned} \tag{6}$$

The probability of being affected is conditional to the presence of the mutation, the probability of which is $P(M|\overline{A}_t)$. Therefore, from equations 2 and 6, we get

$$P(A_{t+n}|\overline{A}_t) = P(M|\overline{A}_t)P_M(A_{t+n}|\overline{A}_t) \tag{7}$$

So

$$P(A_{t+n}|\overline{A}_t) = \left(\frac{1-\pi_t}{2-\pi_t}\right)\left(\frac{\pi_{t+n}-\pi_t}{1-\pi_t}\right) = \frac{\pi_{t+n}-\pi_t}{2-\pi_t} \tag{8}$$

As $\pi_{t+n}$ tends towards 1 when $n$ increases, this probability tends towards $\dfrac{1-\pi_t}{2-\pi_t}$, *i.e.* towards $P(M|\overline{A}_t)$, which is consistent: the probability of eventually being affected is equal to the probability of being a carrier of the mutation because the penetrance tends towards 1.

#### 2.2 Case 2: There is a proportion $x$ of non-penetrant carriers

If a proportion $x$ of individuals are non-penetrant carriers, we have (see equation 5)

$$P_{Mx}(A_{t+n}|\overline{A}_t) = 1 - \frac{P_{Mx}(\overline{A}_{t+n})}{P_{Mx}(\overline{A}_t)}$$

where $P_{Mx}(\overline{A}_{t+n})$ is the probability of being unaffected at age $t+n$. There are two ways to be unaffected: either being in the fraction $x$ of the population or being in the fraction $(1-x)$ while remaining unaffected. So

$$P_{Mx}(\overline{A}_{t+n}) = x + (1-x)(1-\pi_{t+n}) = 1 - (1-x)\pi_{t+n}$$

Similarly

$$P_{Mx}(\overline{A}_t) = x + (1-x)(1-\pi_t) = 1 - (1-x)\pi_t$$

So

$$P_{Mx}(A_{t+n}|\overline{A}_t) = 1 - \frac{1-(1-x)\pi_{t+n}}{1-(1-x)\pi_t} = \frac{(1-x)(\pi_{t+n}-\pi_t)}{1-(1-x)\pi_t} = \frac{\pi'_{t+n}-\pi'_t}{1-\pi'_t}$$

Therefore

$$\begin{aligned} P_x(A_{t+n}|\overline{A}_t) &= P_x(M|\overline{A}_t)P_M(A_{t+n}|\overline{A}_t) \\ &= \left(\frac{1-\pi'_t}{2-\pi'_t}\right)\left(\frac{\pi'_{t+n}-\pi'_t}{1-\pi'_t}\right) \\ &= \frac{\pi'_{t+n}-\pi'_t}{2-\pi'_t} \end{aligned} \tag{9}$$

$\pi_{t+n}$ tends towards 1 with age, so $P_x(A_{t+n}|\overline{A}_t)$ tends towards $\frac{(1-x)-\pi'_{t+n}}{2-\pi'_t}$, which is smaller than $P_x(M|\overline{A}_t)$. Indeed, all the carriers will not become affected because $x > 0$.

### 3 Probability of carrying the mutation for a grandchild of the carrier

The developments above considered the case of the children of the individual carrying the variant. By applying the same approach, we can also compute these probabilities for the grandchildren of the carrier. The age of the child and the grandchildren are noted $t_1$ and $t_2$, respectively.

#### 3.1 Case 1: The penetrance tends towards 1 with age

Following equation 1, the probability of carrying the mutation for a grandchild of the carrier is:

$$P(M|\overline{A}_{t_2}) = \frac{P(\overline{A}_{t_2}|M)P(M)}{P(\overline{A}_{t_2})} \tag{10}$$

where

- $P(M)$ is the probability that the grandchild has inherited the mutation from its parent. $P(M) = \frac{P(M|\overline{A}_{t_1})}{2}$
- $P(\overline{A}_{t_2}|M) = 1-\pi_{t_2}$
- $P(\overline{A}_{t_2}) = 1 - P(A_{t_2})$. $P(A_{t_2})$, the probability of being affected at age $t_2$, is the product of the probability of carrying the mutation, $\frac{P(M|\overline{A}_t)}{2}$, and of the penetrance $\pi_{t_2}$. So $P(\overline{A}_{t_2}) = 1 - \frac{P(M|\overline{A}_t)}{2}\pi_{t_2}$.

Therefore

$$P(M|\overline{A}_{t_2}) = \frac{1-\pi_{t_2}}{\frac{2}{P(M|\overline{A}_{t_1})}-\pi_{t_2}} = \frac{1-\pi_{t_2}}{2\left(\frac{2-\pi_{t_1}}{1-\pi_{t_1}}\right)-\pi_{t_2}} \tag{11}$$

This probability decreases with both the age of the grandchild and the age of its at-risk parent.

#### 3.2 Case 2: There is a proportion $x$ of non-penetrant carriers

With a proportion $x$ of non-penetrant carriers in the population, the probability of carrying the mutation for a grandchild is:

$$P_x(M|\overline{A}_{t_2}) = \frac{P_x(\overline{A}_{t_2}|M)P_x(M)}{P_x(\overline{A}_{t_2})} \tag{12}$$

where

- $P_x(M)$ is the probability that the grandchild has inherited the mutation from its parent. From equation 4, we have: $P_x(M) = \dfrac{P_x(M|\overline{A}_{t_1})}{2} = \dfrac{1-\pi'_{t_1}}{2(2-\pi'_{t_1})}$

- $P_x(\overline{A}_{t_2}|M)$, the probability of not being affected knowing that the mutation is present, is the sum of the probabilities of being non-penetrant, $x$, and of being penetrant and non affected $(1-x)(1-\pi_{t_2})$. So $P_x(\overline{A}_{t_2}|M) = x + (1-x)(1-\pi_{t_2})$.

- $P_x(\overline{A}_{t_2}) = 1 - P_x(A_{t_2})$. $P_x(A_{t_2})$, the probability of being affected at age $t_2$, is the product of three probabilities: (i) carrying the mutation, $\dfrac{P_x(M|\overline{A}_t)}{2}$; (ii) not to belong to the non-penetrant carriers, $(1-x)$; (iii) being affected, which is penetrance $\pi_{t_2}$. So

$$P_x(\overline{A}_{t_2}) = 1 - \frac{P_x(M|\overline{A}_t)}{2}(1-x)\pi_{t_2}$$

Therefore

$$\begin{aligned}
P_x(M|\overline{A}_{t_2}) &= \frac{\dfrac{1-(1-x)\pi_{t_1}}{2(2-(1-x)\pi_{t_1})}(x+(1-x)(1-\pi_{t_2}))}{1-\dfrac{(1-(1-x)\pi_{t_1})(1-x)\pi_{t_2}}{2(2-(1-x)\pi_{t_1})}} \\
&= \frac{(1-\pi'_{t_2})}{2\left(\dfrac{2-\pi'_{t_1}}{1-\pi'_{t_1}}\right)-\pi'_{t_2}} \qquad (13)
\end{aligned}$$

This equation is similar to equation 11, with penetrances multiplied by $(1-x)$.

### 4 Probability of developing the disease later in life for a grandchild of the carrier

#### 4.1 Case 1: The penetrance tends towards 1 with age

If $x = 0$, we can use equation 7

$$P(A_{t_2+n}|\overline{A}_{t_2}) = P(M|\overline{A}_{t_2})P_M(A_{t_2+n}|\overline{A}_{t_2}) \qquad (14)$$

and from equations 6 and 11,

$$P(A_{t_2+n}|\overline{A}_{t_2}) = \frac{1-\pi_{t_2}}{2\left(\dfrac{2-\pi_{t_1}}{1-\pi_{t_1}}\right)-\pi_{t_2}}\left(\frac{\pi_{t_2+n}-\pi_{t_2}}{1-\pi_{t_2}}\right) = \frac{\pi_{t_2+n}-\pi_{t_2}}{2\left(\dfrac{2-\pi_{t_1}}{1-\pi_{t_1}}\right)-\pi_{t_2}} \qquad (15)$$

As $\pi_{t_2+n}$ tends towards 1 with $n$, $P(A_{t_2+n}|\overline{A}_{t_2})$ tends towards $P(M|\overline{A}_{t_2})$. This is expected because in the absence of non-penetrant carriers, all carriers will eventually be affected.

#### 4.2 Case 2: There is a proportion $x$ of non-penetrant carriers

If a proportion $x$ of individuals are non-penetrant carriers, we have (see equation 5)

$$P_{Mx}(A_{t_2+n}|\overline{A}_{t_2}) = 1 - \frac{P_{Mx}(\overline{A}_{t_2+n})}{P_{Mx}(\overline{A}_{t_2})}$$

where

$$P_{Mx}(\overline{A}_{t_2+n}) = x + (1-x)(1-\pi_{t_2+n}) = 1-(1-x)\pi_{t_2+n}$$

and

$$P_{Mx}(\overline{A}_{t_2}) = x + (1-x)(1-\pi_{t_2}) = 1-(1-x)\pi_{t_2}$$

So

$$P_{Mx}(A_{t_2+n}|\overline{A}_{t_2}) = 1 - \frac{1-(1-x)\pi_{t_2+n}}{1-(1-x)\pi_{t_2}} = \frac{(1-x)(\pi_{t_2+n}-\pi_{t_2})}{1-(1-x)\pi_{t_2}} = \frac{\pi'_{t_2+n}-\pi'_{t_2}}{1-\pi'_{t_2}}$$

Therefore,

$$\begin{aligned}
P_x(A_{t_2+n}|\overline{A}_{t_2}) &= P_x(M|\overline{A}_{t_2})P_{Mx}(A_{t_2+n}|\overline{A}_{t_2}) \\
&= \frac{1-\pi'_{t_2}}{2\left(\frac{2-\pi'_{t_1}}{1-\pi'_{t_1}}\right)-\pi'_{t_2}}\left(\frac{\pi'_{t_2+n}-\pi'_{t_2}}{1-\pi'_{t_2}}\right) \\
&= \frac{\pi'_{t_2+n}-\pi'_{t_2}}{2\left(\frac{2-\pi'_{t_1}}{1-\pi'_{t_1}}\right)-\pi'_{t_2}} \qquad (16)
\end{aligned}$$

$\pi_{t+n}$ tends towards 1 with age, so $P_x(A_{t+n}|\overline{A}_t)$ tends towards $\frac{(1-x)-\pi'_{t+n}}{2-\pi'_t}$, which is smaller than $P_x(M|\overline{A}_t)$. Indeed, all the carriers will not become affected because $x>0$.

Making $x$ apparent, the function 16 writes

$$f(x) = \frac{(1-x)(\pi_{t_2+n}-\pi_{t_2})}{2\left(\frac{2-(1-x)\pi_{t_1}}{1-(1-x)\pi_{t_1}}\right)-(1-x)\pi_{t_2}}$$

The second derivative of this function with respect to $x$ is strictly negative in the interval considered, so the function has a maximum.

The first derivative is

$$f'(x) = \frac{\left(\frac{2(1-x)\pi_{t_1}}{(1-(1-x)\pi_{t_1})^2} - 2\left(\frac{2-(1-x)\pi_{t_1}}{1-(1-x)\pi_{t_1}}\right)\right)(\pi_{t_2+n}-\pi_{t_2})}{\left(2\left(\frac{2-(1-x)\pi_{t_1}}{1-(1-x)\pi_{t_1}}\right)-(1-x)\pi_{t_2}\right)^2}$$

The value of $x$ that cancels out $f'(x)$ is solution to the equation

$$\frac{2(1-x)\pi_{t_1}}{(1-(1-x)\pi_{t_1})^2} - 2\left(\frac{2-(1-x)\pi_{t_1}}{1-(1-x)\pi_{t_1}}\right) = 0$$

or, after rearrangements

$$\pi_{t_1}^2 x^2 + (-2\pi_{t_1}^2 + 4\pi_{t_1})x + (\pi_{t_1}^2 - 4\pi_{t_1} + 2) = 0$$

The solution to this quadradic equation for $x \in [0,1]$ is

$$x_m = \frac{-2+\sqrt{2}+\pi_{t_1}}{\pi_{t_1}}$$

Thus, the fraction $x_m$ of non-penetrant carriers for which the probability of declaring the disease in the next $n$ years is maximum depends only on the age-dependent penetrance $\pi_{t_1}$ of the child: the value of $n$ and the age-dependent penetrance of the grandchild play no role.

From the previous equation, we see that the maximum probability of reporting the disease is at $x_m = 0$ if the age-dependence penetrance of the child is

$$\pi_{t_1} \leq 2 - \sqrt{2} \approx 0.586$$

If $\pi_{t_1} > 2 - \sqrt{2}$, $x_m \in ]0, 1]$. As $\pi_{t_1}$ cannot exceed 1, the probability of reporting the disease increases up to $x_m = -1 + \sqrt{2} \approx 0.41$, then tends to 0 as $x$ tends towards 1.
